# Efficacy of Transposition of Plicated Medial Rectus Muscles in the Treatment of A- or V- Pattern Exotropia (An Egyptian Comparative Study)

**DOI:** 10.1101/2024.01.24.24301578

**Authors:** Lamiaa A. El-aidy, Yasser M. Ibrahim, Mohamed A. Elmarakby, Manar A. Ghali

## Abstract

**Objective:** To report effectiveness of bimedial plication and vertical transposition for correction of exotropia associated with A- or V-pattern and compare it with bimedial resection and vertical transposition.

**Methods and analysis:** We retrospectively reviewed the results of surgery of patients who underwent bimedial plication (group I) versus bimedial resection (group II) with vertical offset to correct both exotropia and pattern deviation not secondary to oblique overaction in the period between January 2021 and January 2023. Results of both groups were compared. Success was considered when pattern deviation was ≤ 8 prism diopters PD and horizontal deviation was within 10 PD of orthophoria at 6 months postoperatively.

**Results:** The mean preoperative angle of exotropia in PD was 40.6 ± 7.2 in group I and 41.1 ± 7.5 in group II (p=.8). At 6 months postoperatively, they achieved angle of 4.6 ± 4.1 and 2.5 ± 4.5 PD respectively (no significant difference between both groups; p=.19). The mean preoperative pattern deviation was 21.3 ± 3.7 PD in group I and 21.6 ± 4.4 in group II. Postoperative pattern collapse was 16.7 ± 5.6 and 16.6 ± 4.1 respectively. The percentage of success in pattern collapse was 90.6% in group I and 84.4% in group II (no significant difference between both groups; p=.71).

**Conclusion:** Vertical transposition with plication of medial recti is a safe, effective, and rapid technique for correction of cases of A- or V- pattern exotropia not associated with oblique overaction. Results were not significantly different from resection/vertical transposition.

**What is already known in this topic?:**

- A- or V-pattern exotropia without significant inferior oblique overaction can be treated by vertical transposition of the horizontal recti after ordinary recession or resection.
- Some studies reported muscle plication and transposition as an alternative technique to resection and transposition. Plication allow preservation of ciliary circulation and lowering the incidence of muscle loss.

**What this study adds?:**

- In this retrospective study, we concluded that medial rectus plication and transposition is an effective alternative technique to medial rectus resection and transposition in treatment of A- or V- pattern exotropia without significant inferior oblique overaction (No significant difference between both techniques).

**What this study might affect research, practice or policy?:**

- Vertical Transposition of plicated medial recti is a new promising technique which should be considered in managing cases of A- or V-pattern exotropia without significant inferior oblique overaction.

## Introduction

The amount of horizontal deviation in strabismic patients may differ in upgaze and downgaze apart from the primary position. This difference is called A- and V-patterns or alphabetical patterns [1].

A-pattern is described when the difference in horizontal deviation between upgaze and downgaze is more than 10 PD, with more divergence in downgaze, while V-pattern is described when this difference is more than 15 PD, with more divergence in upgaze [2].

The reported prevalence of A- or V-patterns in patients with horizontal deviation ranges between 20% and 50% in different studies [3].

Many theories were proposed to clarify the etiology of alphabetical patterns, like: oblique muscle overaction or palsy, nerve misdirection such as Duane syndrome, orbital factors as craniofacial abnormalities, heterotopia of muscle pulleys, extraocular muscles’ anomalies, and disruption of fusion [4,5].

Intermittent exotropia is the most common type of exodeviation. It is a large phoria that can be controlled intermittently by fusional mechanisms and may later progress spontaneously into constant exotropia with loss of control. A- and V-patterns are seen in a large number of patients with typical intermittent exotropia [1,7].

Detection of pattern deviation in cases of exotropia or horizontal deviation — in general — is very important before determining surgical intervention. Unrecognized patterns may lead to either postoperative over or undercorrection or may raise the need for more surgeries to improve the undesirable surgical outcomes [2].

The generally-accepted surgical corrections of pattern strabismus include oblique muscle weakening when there is obvious oblique muscle overaction [9,10], and horizontal muscle transposition when there is not [11–13].

However, both procedures can be done together in large patterns [2].

Other approaches include surgery on vertical recti (nasal or temporal horizontal displacement of insertions) and slanting of horizontal recti insertions [12].

Many options are advocated for the correction of exotropia: unilateral recession resection, bilateral lateral rectus recession or bimedial resection. A three or four-muscle surgery may be done in large angle exotropia [13].

Muscle plication is an alternative surgical procedure to muscle resection, used to strengthen the muscle. It has the benefit of being potentially reversible as well as being better in preserving the anterior ciliary circulation; thus lowering the risk of anterior segment ischemia [16,17].

Plication also reduces the risk of a lost muscle, and it can be performed using adjustable-suture techniques [16].

The combination of both vertical transposition and plication of the horizontal recti is one of the newer techniques used for the correction of cases with both vertical and horizontal strabismus (as elaborated in the discussion) [14,16].

In this retrospective cohort study, we chose cases with primary exotropia — either constant or intermittent — associated with pattern deviation who underwent either medial rectus muscle plication and transposition or the standard technique in which resection is done instead of plication with either superior or inferior transposition according to type of pattern (A or V). We hypothesized that this technique is as effective as bimedial resection and transposition. However, bimedial plication and transposition will offer advantages regarding safety, short operation time as well as decreasing incidence of postoperative anterior segment ischemia.

### Subjects and methods

The study was approved by the Zagazig University Institutional Review Board (IRB). Informed consent was taken from patients or the parents (for children less than 16 years old). All surgeries were performed at Ophthalmology department of Zagazig University Hospitals.

The medical records of patients diagnosed with A- or V- pattern exotropia with no or minimal oblique overaction, who underwent strabismus surgery between January 2021 and January 2023; were retrospectively reviewed. Only patients with primary exotropia (either constant or intermittent) with angle of deviation ≥ 30 and ≤ 55 PD; were included. A-pattern exotropia was included when the difference in exodeviation between upgaze and downgaze was more than 10Δ and V-pattern when this difference was more than 15Δ. Age ranged from 4 to 50 years old.

Patients with oblique muscle overaction (≥ +2), sensory exodeviation, anisometropia ≥5 diopters, axial length < 22.5 or > 25 mm, associated dissociated vertical deviation DVD, or less than 6 months’ follow-up were excluded.

Preoperative data collected included best corrected visual acuity in Snellen decimal, cycloplegic refraction (using 1% cyclopentolate chloride) with retinoscopy or autorefractometry, extraocular muscle movement, degree of ocular deviation in primary position at distance (6 meters) and at near (1/3 meter) by prism and alternating cover test, horizontal deviation at 1/3 meter (in upgaze and downgaze, about 25 degrees up and down respectively without glasses to avoid prismatic effect of spectacle correction), pattern deviation is the difference in horizontal deviation between up and downgazes, oblique muscle overaction grading from +1 to +4 [17], stereoacuity at near by Titmus test (with poor being >200 arcsec and moderate being between 80 and 100), slit lamp examination of the anterior segment and fundus examination by indirect ophthalmoscope.

After matching all preoperative data based on age (in years), sex, BCVA, refraction, preoperative angle of exotropia, V pattern and A pattern, we divided patients in two groups, each group was further divided into 2 subgroups A and V referring to A- and V-patterns: 1) Group I (32 patients) underwent bilateral medial rectus plication and vertical transposition, 2) Group II (control group, 32 patients) underwent bimedial resection and vertical transposition. The amount of A- and V- pattern collapse (the difference between pre and postoperative pattern deviation) was calculated at 1 month and 6 months postoperatively. Success was considered when pattern deviation was ≤8 PD at 6 months postoperatively. Success in horizontal deviation was considered when deviation was 10 PD within orthophoria at 6 months postoperatively.

### Surgical technique

All surgical procedures were performed under general anesthesia by the same surgeon (MG). A limbal-conjunctival incision was done. It’s about three hours length with 2 horizontal releases at the end of the incision to make the opening wider. The medial rectus muscle was exposed and freed from the Tenon’s tissue to the planned distance for resection or plication.

For the plication (Figure 1), one double-armed 6-0 Vicryl suture was passed through the center and both muscle edges at the desired distance from the muscle’s insertion and locked. Then, each suture was placed toward the site of transposition. For example, if we targeted to transpose the medial rectus a full tendon width superiorly, the inferior suture was passed under the medial rectus to the superior edge of muscle insertion. The superior suture was passed partial thickness through the sclera, just anterior to the muscle insertion, with a vertical offset about 8-10 mm. Then, the inferior suture was passed through the sclera approximately at the superior edge of the insertion. Both sutures were tied together which resulted in a triangular fold in the muscle.

**Figure 1.** Medial rectus muscle plication and vertical transposition. **a** The medial rectus is stretched between the two muscle hooks. **b** A single double-armed Vicryl suture was passed through the muscle center to the superior edge. **c** The suture was passed to the inferior edge after locking the superior one. **d** Picture after locking both sutures. **e** Inferior suture was passed through the sclera at the superior pole of muscle insertion and the superior suture was passed through the sclera a full-tendon-width above the first scleral path. **f** Position of the muscle after tying the sutures. **g** Final appearance of the folded and transposed muscle.

For the resection, two single-armed Vicryl sutures were passed through both muscle edges at the desired distance from the muscle insertion and tied, then the muscle was disinserted from the globe and resected just anterior to the previously-passed sutures. The muscle was then transposed and secured to the sclera at the desired position with partial-thickness scleral passes.

In the A-pattern group, medial recti were displaced upward (toward the apex of the pattern) by 5 mm if the difference between up and downgazes was 10- ≤ 20 PD and 8-10 mm if > 20 PD. In the V-pattern group, they were displaced downward by 5 mm if the difference between up and downgazes was 15-≤ 25 PD and 8-10 mm if the difference > 25 PD.

The amount of horizontal muscle operation was done according to Kenneth Wright’s tables for strabismus [18]. We decreased amount of resection/plication by 1 mm according to surgeon’s experience.

### Statistical analysis

The collected data was analyzed using the Statistical Package for the Social Sciences (SPSS) version 28 (IBM Corp., Armonk, NY, USA). Data was double checked for normality using normality plots and Shapiro-Wilk test. Data was summarized using mean and standard deviation in quantitative data and using frequency (count) and relative frequency (percentage) for categorical data. Comparisons between groups were done using unpaired t-test in normally distributed quantitative variables while non-parametric Mann-Whitney test was used for non-normally distributed quantitative variables. Comparison between pre-and postoperative values was done using paired t-test in normally distributed quantitative variables while non-parametric Friedman test and Wilcoxon signed rank test were used for non-normally distributed quantitative variable [19]. For comparing categorical data, Chi square (χ2) test was performed. Exact test was used instead when the expected frequency was less than 5 [20]. p-values less than 0.05 were considered significant.

## Results

64 patients were reviewed, 32 were treated with bilateral medial rectus plication and vertical transposition (Group I) and 32 patients treated with bilateral medial rectus resection and vertical transposition (Group II). Each group was further divided into two subgroups: A and V. The mean age of all patients at the time of operation was 15.95 ± 12.79 years (range: 4-50). Table 1 shows the preoperative criteria in both groups (no significant difference between both groups).

**Table 1:**
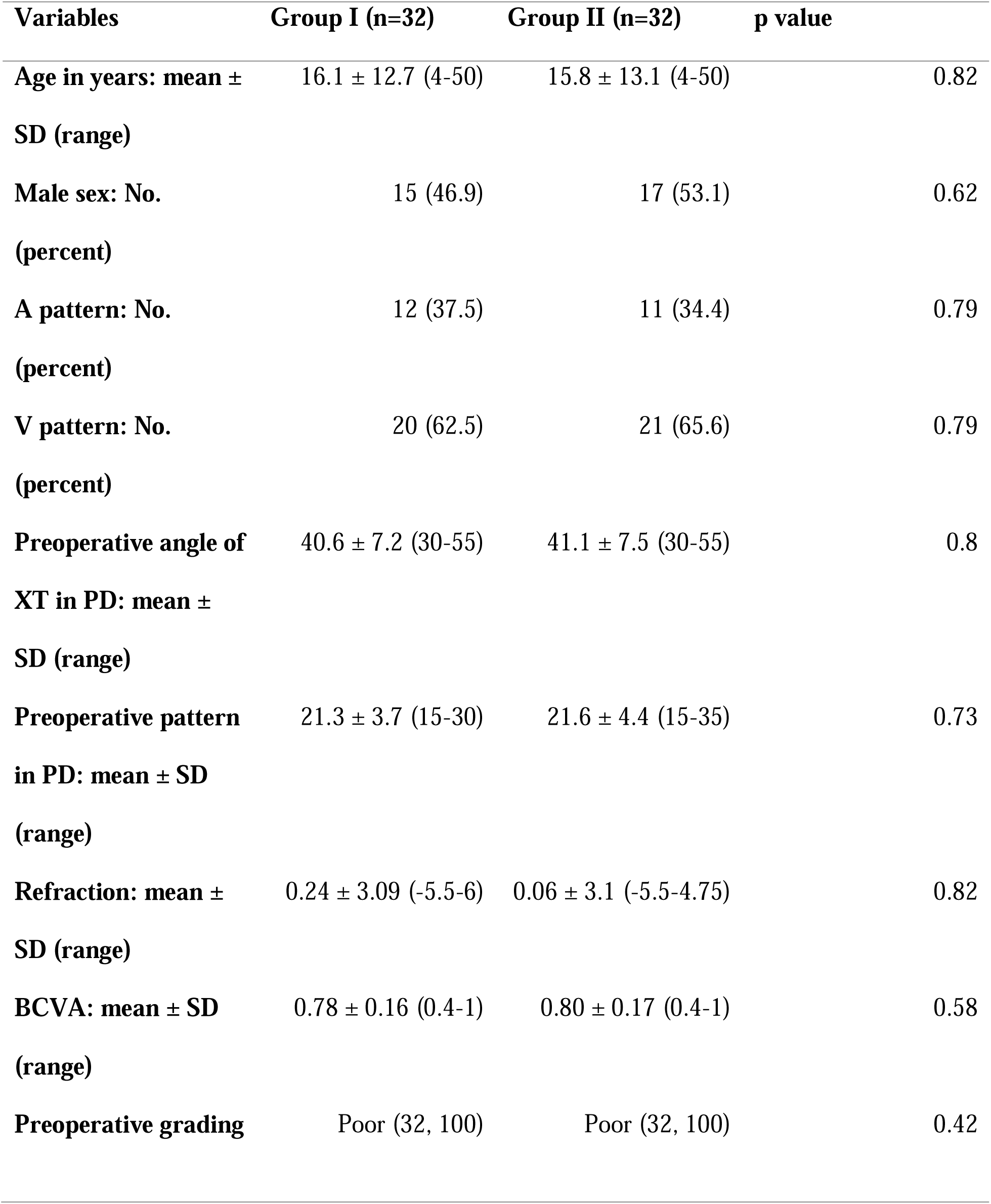
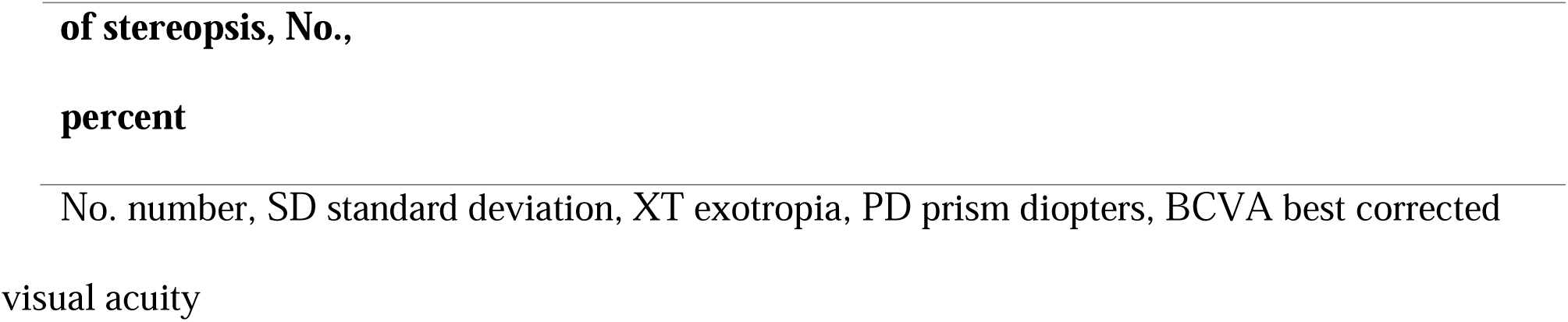
Preoperative data for the studied groups.

| **Variables** | **Group I (n=32)** | **Group II (n=32)** | **p value** |
| --- | --- | --- | --- |
| **Age in years: mean ± SD (range)** | 16.1 ± 12.7 (4-50) | 15.8 ± 13.1 (4-50) | 0.82 |
| **Male sex: No. (percent)** | 15 (46.9) | 17 (53.1) | 0.62 |
| **A pattern: No. (percent)** | 12 (37.5) | 11 (34.4) | 0.79 |
| **V pattern: No. (percent)** | 20 (62.5) | 21 (65.6) | 0.79 |
| **Preoperative angle of XT in PD: mean ± SD (range)** | 40.6 ± 7.2 (30-55) | 41.1 ± 7.5 (30-55) | 0.8 |
| **Preoperative pattern in PD: mean ± SD (range)** | 21.3 ± 3.7 (15-30) | 21.6 ± 4.4 (15-35) | 0.73 |
| **Refraction: mean ± SD (range)** | 0.24 ± 3.09 (-5.5-6) | 0.06 ± 3.1 (-5.5-4.75) | 0.82 |
| **BCVA: mean ± SD (range)** | 0.78 ± 0.16 (0.4-1) | 0.80 ± 0.17 (0.4-1) | 0.58 |
| **Preoperative grading** | Poor (32, 100) | Poor (32, 100) | 0.42 |

Table 2 shows the intraoperative data and the postoperative outcomes in both groups. Postoperative angles at 6 months were 4.6 ±6.0 and 2.53 ±6.5 PD exodeviation respectively (no significant difference in both groups regarding postoperative horizontal deviation, p= 0.185). No significant difference between both groups was detected regarding stereopsis as a sensory outcome.

**Table 2:** Intraoperative and postoperative data in both groups.

| **Variables** | **Group I**<br>**(n= 32)** | **Group II**<br>**(n= 32)** | **p value** |
| --- | --- | --- | --- |
| **Amount of MR**<br>**plication/resection,**<br>**mm, mean ± SD**<br>**(range)** | 4.6 ± 0.8 (3-6) | 4.6 ± 0.9 (3-6.5) | 0.87 |
| **Amount of MR**<br>**transposition, mm,**<br>**mean ± SD (range)** | 6.1 ± 2.1 (5-10) | 6.1 ± 2.1 (5-10) | 1 |
| **Postoperative XT at 1**<br>**m, PD, mean ± SD**<br>**(range)** | -1.4 ± 6.6 (-25 to 10) * | -1.8 ± 6.5 (-25 to 10) | 0.33 |
| **Postoperative XT at 6**<br>**m, PD, mean ± SD**<br>**(range)** | 4.6 ± 6.0 (-12 to 20) | 2.5 ± 6.5 (-15 to 20) | 0.19 |
| **Pattern collapse** | 16.7 ± 5.6 | 16.1 ± 4.1 | 0.65 |
| **Postoperative grading**<br>**of stereopsis, No.,**<br>**percent** | Poor (7, 21.9)<br>Moderate (24, 75)<br>Good (1, 3.1) | Poor (6, 18.8)<br>Moderate (25, 78.1)<br>Good (1, 3.1) | 0.56 |

MR medial rectus, XT exotropia, \* minus sign refers to esotropia and vice versa, No. number.

Regarding pattern deviation, the mean preoperative pattern deviation (difference in prism diopters between up and down gazes) was 21.3 ± 3.7 PD in group I, with a postoperative pattern collapse of 16.7 ± 5.6 PD. In group II, the mean preoperative pattern deviation was 21.6 ± 4.4 PD, with a postoperative pattern collapse of 16.1 ±4.1 PD. Change between pre and postoperative pattern deviation was significant in each group. The percentage of success in pattern collapse (pattern deviation ≤ 8 PD) was 90.6% in group I and 84.4% in group II. There was no significant difference in success between both groups (p=.708). There were no intra or postoperative complications recorded.

The results of the subgroups regarding pattern deviation are shown in table 3, which shows significant collapse in pattern deviation postoperatively in the four subgroups.

**Table 3:**
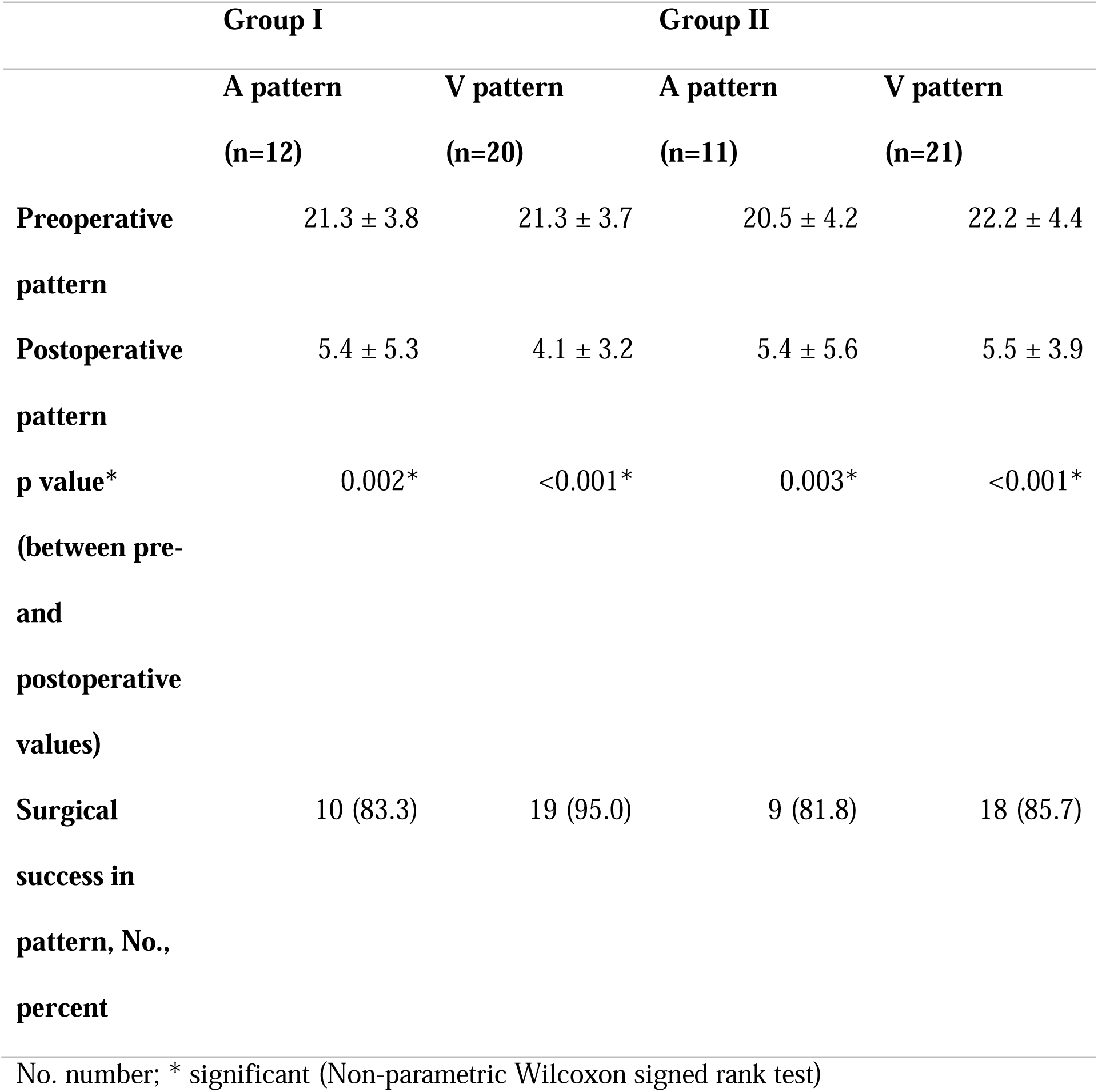
Preoperative pattern and postoperative surgical correction in subgroups (Mean ±SD)

## Discussion

Pattern deviation can be corrected either by oblique muscle surgery in cases with significant oblique muscle overaction or with horizontal muscles transposition in cases without oblique overaction.

In this retrospective study, we chose only cases with primary exotropia either intermittent or constant with no oblique overaction. We compared the effect of bimedial plication and transposition as an alternative technique with bimedial resection and transposition as the standard.

Muscle transposition was prescribed in literature either by doing complete muscle disinsertion as in the Knapp procedure or without muscle disinsertion as in the Nishida procedure. However, both don’t have a strengthening effect; they aimed only to change the pulling force of the muscle [12].

**Gokyigit B et al** [14,23] were the first to present this technique (rectus plication and transposition) and named it “sliding shape extraocular muscle transposition with plication procedure (SSMTP)”. There were 10 cases in their study (out of 17 cases) with exotropia and vertical deviation, 5 of them were treated by medial rectus plication and transposition as a primary surgical intervention except one case who had done one surgery before. There was no anterior segment ischemia even in cases who had done 3 recti muscles surgery on the same eye. Of ten cases of exotropia, five achieved pattern deviation of less than or equal 8 PD, and three achieved horizontal deviation within 10 PD of orthophoria. They prescribed this technique to be safe, effective, and with good results on long-term follow-up (between 24 and 40 months). Our study differed from **Gokyigit B et al** in that our sample size was larger (32 patients) and we chose only cases of primary exotropia. However, the follow up period was shorter (6 months).

Additionally, **Shah PR& Pihlblad MS** [14] published a report of 4 successful cases of complex horizontal and vertical squint, who were treated with unilateral recession/plication with vertical transposition. The technique was effective, and all four cases achieved the required ocular alignment for both horizontal and vertical deviations. No ischemia occurred even in cases with previous squint surgery in the same eye.

In our study, both horizontal and vertical successes were achieved in both groups. Success in horizontal deviation was defined as being within 10 PD of orthophoria at 6 months postoperatively. The percentage of success in group I was 81.3% (26 out of 32 patients) and in group II was 84.4% (27 out of 32), no significant difference between both groups, p=0.74. **ElKamshoushy A** [22] also reported good results with bimedial resection in cases of primary exotropia, success rate was 77%. However, the study included only cases with large angle exotropia.

According to the surgical design in group I, we used a limbal approach and a double armed 6/0 Vicryl suture which passed from the muscle center to the periphery in both sides and was tied. The two sutures were then tied after passing through partial scleral thickness.

**In Shah and Pihlblad’s** study, fornix approach was used and two separate single-armed polyglactin sutures were passed at the muscle edges and tied. **Gokyigit B et al** used limbal approach and either single-or double-armed sutures. In six patients, they added a central 6/0 Dacron suture to make the whole muscle edge at the same plication line to maintain the effect of plication.

We believe that our technique avoids muscle splitting and decreases the need for a central muscle suture to enforce the effect of plication. One disadvantage we noticed was that sheath wiring of the sclera may occur after tying the sutures if the scleral paths are too superficial. This can be avoided by proper passage through the sclera and avoiding superficial paths.

Our results in plication and resection were comparable with no significant difference between both groups. This is like many studies which recorded similar results between plication and resection [23–25].

Anterior segment ischemia is a rare complication of strabismus surgery which occurs secondary to muscle disinsertion either by recession or resection. The risk increases if three muscles are operated on in the same eye. Plication offers a good alternative to muscle resection with preservation of anterior ciliary circulation especially in surgeries where 3-muscle surgery is needed [24].

The unsuccessful cases in pattern collapse were 3 (9.4%) in group I and 5 (15.6%) in group II (p=.708). The causes of residual pattern may be related to unrecognized oblique overaction, inaccurate measurements of pattern preoperatively, or improper execution of the surgical technique.

The age, demographic and preoperative data were comparable with no significant differences in both groups. Other strength points in our study were: the presence of a control group and the average follow-up period of 6 months.

However, a larger sample is needed, and a longer follow-up period is recommended to address the long-term effects of the new technique.

In conclusion, we find that horizontal transposition of plicated medial rectus muscle is a safe, rapid, effective, and simple technique for the correction of primary exotropic patients with A- or V-pattern not secondary to oblique muscle overaction.

## Data Availability

All data produced in the present study are available upon reasonable request to the authors

## Authorship

All authors meet the International Committee of Medical Journal Editors (ICMJE) criteria for authorship for this manuscript, take responsibility for the integrity of the work as a whole, and have given final approval for this version to be published.

## Compliance with ethics guidelines

All procedures followed were in accordance with the ethical standards of the responsible committee on human experimentation (institutional and national) and with the Helsinki Declaration of 1964, as revised in 2013 Informed consent was obtained from all patients included in the study.

## Author contributions

All authors contributed to the study conception and design. Material preparation, data collection and analysis were performed by Lamiaa El-aidy and Manar Ghaly. The first draft of the manuscript was written by Lamiaa El-aidy and all authors commented on previous versions of the manuscript. All authors read and approved the final manuscript.

## Data availability

The datasets generated during and/ or analyzed during the current study are available from the corresponding author on reasonable request.

## References

[1] K. W. Wright, “Alphabet Patterns and Oblique Muscle Dysfunctions,” in Handbook of Pediatric Strabismus and Amblyopia, K. W. Wright, P. H. Spiegel, and L. S. Thompson, Eds., New York, NY: Springer New York, 2006, pp. 284–322. doi: 10.1007/0-387-27925-3_9.

[2] H. T. Sekeroglu, K. E. Turan, S. Uzun, E. C. Sener, and A. S. Sanac, “Horizontal muscle transposition or oblique muscle weakening for the correction of V pattern?,” Eye, vol. 28, no. 5, pp. 553–556, May 2014, doi: 10.1038/eye.2014.16.

[3] S. M. Straight and R. S. Bahl, “A- and V-Pattern Strabismus,” in Practical Management of Pediatric Ocular Disorders and Strabismus: A Case-based Approach, E. Traboulsi and V. Utz, Eds., New York, NY: Springer New York, 2016, pp. 583– 592. doi: 10.1007/978-1-4939-2745-6_54.

[4] B. J. Kushner, “Effect of ocular torsion on A and V patterns and apparent oblique muscle overaction.,” Arch Ophthalmol, vol. 128, no. 6, pp. 712–718, Jun. 2010, doi: 10.1001/archophthalmol.2010.88.

[5] Y. B. Lee et al., “Effect of horizontal rectus surgery for the correction of intermittent exotropia on sub-A or sub-V pattern.,” PLoS One, vol. 12, no. 6, p. e0179626, 2017, doi: 10.1371/journal.pone.0179626.

[6] K. H. Shin, H. J. Lee, and H. T. Lim, “Ocular torsion among patients with intermittent exotropia: relationships with disease severity factors.,” Am J Ophthalmol, vol. 155, no. 1, pp. 177–182, Jan. 2013, doi: 10.1016/j.ajo.2012.07.011.

[7] K. Kumar, H. N. Prasad, S. Monga, and R. Bhola, “Hang-back recession of inferior oblique muscle in V-pattern strabismus with inferior oblique overaction,” Journal of American Association for Pediatric Ophthalmology and Strabismus, vol. 12, no. 4, pp. 401–404, Aug. 2008, doi: 10.1016/j.jaapos.2008.01.015.

[8] S. Akar, B. Gökyiğit, and O. F. Yilmaz, “Graded anterior transposition of the inferior oblique muscle for V-pattern strabismus.,” J AAPOS, vol. 16, no. 3, pp. 286–290, Jun. 2012, doi: 10.1016/j.jaapos.2012.01.009.

[9] Y. Oya, T. Yagasaki, M. Maeda, M. Tsukui, and K. Ichikawa, “Effects of vertical offsets of the horizontal rectus muscles in V-pattern exotropia without oblique dysfunction.,” J AAPOS, vol. 13, no. 6, pp. 575–577, Dec. 2009, doi: 10.1016/j.jaapos.2009.09.017.

[10] A. M. Mostafa and R. R. Kassem, “A comparative study of medial rectus slanting recession versus recession with downward transposition for correction of V-pattern esotropia.,” J AAPOS, vol. 14, no. 2, pp. 127–131, Apr. 2010, doi: 10.1016/j.jaapos.2009.11.025.

[11] A. Dickmann et al., “Effect of vertical transposition of the medial rectus muscle on primary position alignment in infantile esotropia with A- or V-pattern strabismus.,” J AAPOS, vol. 15, no. 1, pp. 14–16, Feb. 2011, doi: 10.1016/j.jaapos.2010.11.017.

[12] B. Gokyigit, A. Inal, B. Ocak, and E. D. Aygit, “Sliding Shape Extraocular Muscle Transposition with Plication: A Novel Method,” Beyoglu Eye Journal, vol. 4, no. 2, pp. 120–122, 2019.

[13] Z. I. Currie, T. Shipman, and J. P. Burke, “Surgical correction of large-angle exotropia in adults.,” Eye (Lond), vol. 17, no. 3, pp. 334–339, Apr. 2003, doi: 10.1038/sj.eye.6700347.

[14] P. R. Shah and M. S. Pihlblad, “Transposition of plicated horizontal muscles,” Journal of American Association for Pediatric Ophthalmology and Strabismus, vol. 24, no. 4, pp. 244–247, Aug. 2020, doi: 10.1016/j.jaapos.2020.04.008.

[15] K. W. Wright, “Rectus Muscle Plication Procedure,” JAMA Ophthalmology, vol. 133, no. 2, pp. 226–227, Feb. 2015, doi: 10.1001/jamaophthalmol.2014.4259.

[16] B. Gokyigit et al., “Long-term results of ‘sliding shape muscle transposition with plication’ operation,” Journal of American Association for Pediatric Ophthalmology and Strabismus, vol. 23, no. 4, p. e30, Aug. 2019, doi: 10.1016/j.jaapos.2019.08.106.

[17] K. W. Wright and Y. N. J. Strube, “Inferior Oblique Muscle Weakening Procedures,” in Color Atlas Of Strabismus Surgery: Strategies and Techniques, Fourth edition., K. W. Wright and Y. N. J. Strube, Eds., New York, NY: Springer New York, 2015, pp. 135–136. doi: 10.1007/978-1-4939-1480-7_5.

[18] K. W. Wright and Y. N. J. Strube, “Appendix A: Surgical numbers,” in Color Atlas Of Strabismus Surgery: Strategies and Techniques, Fourth edition., K. W. Wright and Y. N. J. Strube, Eds., New York, NY: Springer New York, 2015, pp. 191–192. doi: 10.1007/978-1-4939-1480-7_5.

[19] Y. H. Chan, “Biostatistics 102: quantitative data--parametric & non-parametric tests.,” Singapore Med J, vol. 44, no. 8, pp. 391–396, Aug. 2003.

[20] Y. H. Chan, “Biostatistics 103: qualitative data - tests of independence.,” Singapore Med J, vol. 44, no. 10, pp. 498–503, Oct. 2003.

[21] B. Gokyigit, A. İnal, O. B. Ocak, and E. D. Aygıt, “Sliding shape extraocular muscle transposition with plication: Long-term results.,” Int Ophthalmol, vol. 41, no. 11, pp. 3593–3598, Nov. 2021, doi: 10.1007/s10792-021-01932-9.

[22] A. A. ElKamshoushy, “Bilateral medial rectus resection for primary large-angle exotropia.,” J AAPOS, vol. 21, no. 2, pp. 112–116, Apr. 2017, doi: 10.1016/j.jaapos.2017.03.003.

[23] Y. Kimura and T. Kimura, “Comparative study of plication–recession versus resection–recession in unilateral surgery for intermittent exotropia,” Japanese Journal of Ophthalmology, vol. 61, no. 3, pp. 286–291, May 2017, doi: 10.1007/s10384-017-0501-5.

[24] P. A. Huston and D. L. Hoover, “Surgical outcomes following rectus muscle plication versus resection combined with antagonist muscle recession for basic horizontal strabismus.,” J AAPOS, vol. 22, no. 1, pp. 7–11, Feb. 2018, doi: 10.1016/j.jaapos.2017.09.004.

[25] M. Stunkel, A. J. Cantor, and D. A. Plager, “Intermediate-term outcomes of horizontal muscle plication versus resection,” Journal of American Association for Pediatric Ophthalmology and Strabismus, 2021, [Online]. Available: https://api.semanticscholar.org/CorpusID:240618297

